# Maternal delays and associated factors in utilizing institutional delivery in East Wallaga Zone, Oromia, Ethiopia

**DOI:** 10.1101/2025.02.04.25321654

**Authors:** Getahun Tulu Amante, Gemechu Dereje Feyissa, Markos Desalegn, Emiru Merdassa, Mokonnen Dereje

## Abstract

**Background:** Pregnancy and childbirth-related complications are unpredictable; however, they are preventable by timely care-seeking to obstetric care services. Maternal delay in utilizing institutional delivery is associated with high maternal mortality in developing countries, including Ethiopia. There were limited studies on the magnitude and associated factors with maternal delays in the study area.

**Objective:** To assess the magnitude of maternal delays and associated factors in utilizing institutional delivery among women who gave birth at public hospitals in East Wallaga Zone, Oromia, Ethiopia, 2023.

**Methods:** A facility-based cross-sectional study design complemented by the qualitative inquiry was used to conduct the study. The data collection period was from February 4 to April 10, 2023, and 422 systematically selected mothers were included in the study. The data was collected by trained data collectors using an interviewer-administered, pre-tested structured questionnaire, and the collected data was checked for completeness, entered into Epi-Data version 4.6, cleaned, and exported to SPSS version 24.0. The association between dependent and independent variables was estimated by an adjusted odds ratio (AOR) along with a 95% confidence interval (CI). A P-value < 0.05 was considered to declare statistical significance.

**Result:** The study revealed that 47.2% [95% CI: 42.7, 51.7)] of the mothers experienced a first delay, 71.3% [95% CI: 64.5, 78.1)] of them experienced a second delay, and 10.2% [95% CI: 9.23, 11.17)] experienced a third delay. Obstetric complications during the current pregnancy [AOR = 1.73, 95% CI: (1.08, 2.76)] and referral cases [AOR = 1.60, 95% CI: (1.04, 2.47)] were associated with the first delay. Poor knowledge of danger signs of labor [AOR = 2.93, 95% CI: 1.47, 5.86] was associated with the second delay. Obstetric complications during the current pregnancy [AOR = 2.26, 95% CI: (1.01, 5.05)] and lack of money for transportation [AOR = 3.98, 95% CI: (1.66, 9.57)] were associated with the third maternal delay in utilizing institutional delivery.

**Conclusion:** The current study showed that the magnitude of first- and second-maternal delays in the utilization of institutional delivery services was high. Obstetric complications during current pregnancy, referral cases, poor knowledge of danger signs of labor, and lack of money for transportation were associated with maternal delays in utilizing institutional delivery. To further reduce delays, this study emphasizes the significance of addressing three delays in seeking institutional delivery services. The government’s support is indispensable to help the mothers in prompt utilization of institutional delivery, especially in supporting the referral linkages as more mothers encounter delays after they had been referred from the first institution. Making service free of charge only cannot reduce maternal delays by itself, but the provision of free ambulance services must be strengthened to accompany the mothers to the health institutions. When the need arises for the mothers to be provided with free service, they should first be accompanied to the health facilities; otherwise, the maternal delays can even endanger the lives of the mothers or their newborns.

## Introduction

Pregnancy and childbirth are critical periods in women’s life. Timely and appropriate utilization of institutional delivery services is mandatory as the health of women and their newborns in developing countries can be jeopardized by pregnancy and childbirth-related complications (1). Delay for utilizing institutional delivery refers to the time interval from deciding to seek emergency care to start receiving of first healthcare. Maternal delays in utilizing institutional delivery refer to failure to decide to seek care within one hour, failure to reach the health facility within one hour of making the decision or waiting there more than one hour to receive delivery care, and failure to take responsibility for accompanying the mother to the health facility (2).

According to the World Health Organization (WHO), reducing maternal delays in the utilization of emergency obstetric care is one of the measures implemented to prevent high maternal mortality in developing countries. Serious pregnancy and childbirth-related complications arise unpredictably. A fatal complication may arise among 15% of all pregnant women in the process of childbirth (3). This may require the mothers to get skilled delivery attendance, including specialized obstetrical interventions even to survive (4).

Globally, unpredictable serious complications occur in 10-15% of the 210 million pregnancies each year (5) but, most of them can be managed by increasing access to skilled birth attendance (6). Approximately half a million women die from causes related to the process of childbirth every day worldwide. Of these, almost all (99%) of them are occurring in low-resource settings. Besides, 2.8 million newborns lose their lives annually within 28 days of birth, with 2 million occurring within the early neonatal period and there are 2.6 million stillbirths with 45 % of them occurring during childbirth or labor (3).

From an estimated 2.9 million women giving birth every year, approximately over 25,000 women and girls die each year and more than 500,000 sustain serious injuries with permanent damage to their health, including obstetric fistulas. It is estimated that 100,000 women suffer from untreated fistulas across the country and another 9,000 women develop fistulas every year which are attributable to obstructed labor and a lack of maternal health care (7). This can be halted by increased access to and utilization of emergency obstetric care (8).

Several factors lead to delays in access to and utilization of institutional delivery services such as distance, women’s autonomy, and lack of resources are the main factors associated with delays in access to health care (9). Access to health facilities can be inhibited by different factors including maternal decision-making power, socio-economic and cultural factors, lack of and/or cost of transport, and health system factors (10).

Maternal delays are the major factors that are associated with high indirect maternal mortality (MM) in developing countries including Ethiopia (11). Even though timely utilization of institutional delivery is one of the golden intervention strategies to minimize maternal death; there are still time gaps in using the services differing among and within populations happening during labor and delivery (10). Delays in receiving timely and appropriate care in the event of a pregnancy complication have been put forward as a major determinant in maternal mortality (10, 12).

Progress on this issue will be the ultimate judge of sustainable development. The Sustainable Development Goals (SDGs) with a vision of EPMM by 2030 further call for an acceleration of current progress with the target of achieving a global maternal mortality ratio (MMR) of 70 per 100, 000 live births, or less (13). To catch up with these global aspirations, Ethiopia has currently set a five-year health sector transformation plan (HSTP) to reduce MMR to 199 per 100,000 live births by 2020 and failed to achieve it. According to the 2016 Ethiopian Demographic and Health Survey (EDHS), MMR was estimated to be 412 per 100,000 live births (14). The current discrepancy in maternal mortality rates between developed and developing countries is due to differences in the timely management of obstetric complications (15).

Although maternal health care utilization is essential for further improvement of maternal and child health, little is known about the magnitude of timely use and factors influencing the use of these services in the study area. Hence this study aimed to fill this gap to describe the current status of delays in utilizing institutional delivery among women who gave birth at public hospitals in the East Wallaga Zone, Oromia, Ethiopia.

## Methods

### Study area and period

The study was conducted among four public hospitals selected by convenience including Wallaga University Referral Hospital, Nekemte Comprehensive Specialized Hospital, Sire Primary Hospital, and Arjo Primary Hospital in East Wallaga Zone which is located in Oromia Regional State, western Ethiopia. The Zonal Health Department is located in Nekemte town which is located 331km to the west of Addis Ababa, the capital city of Ethiopia.

As to the projection from the 2007 census, the zone has 1,660,124 total population with 827,899 males and 83,224 females. There are 4 public hospitals, 65 public health centers, and 325 health posts with a total of 2, 254 health workforce. There are 365,228 reproductive-age women and 66,405 of them are pregnant. Annual zonal institutional delivery utilization for the 2014 Ethiopian fiscal year was 47%. The status of timely utilization of institutional delivery is not known in the study area. The data collection period was from February 4 to April 10, 2023.

### Study design, study population, and inclusion criteria

A facility-based cross-sectional study design was employed to conduct the study. All women using institutional delivery at public hospitals in the East Wallaga zone were the source population. Systematically selected women using institutional delivery at selected public hospitals during a study period and who fulfilled the inclusion criteria were considered for the study. The inclusion criteria were all women getting delivery service in the selected public hospitals in the East Wallaga zone at the period of data collection, excluding women who were admitted with obstetric complications before the onset of labor and those who did not get verbal consent to participate in the study.

### Sample size determination

The sample size for the study was calculated by using a single population proportion formula by using the prevalence of a maternal delay from a similar study done in Jimma Medical Center which identified the prevalence of maternal delays in utilizing institutional delivery service was 46.7% (31).

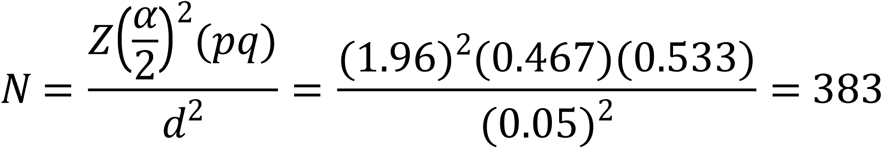

z = the standard normal deviate, usually set at 1.96 which corresponds to a 95% confidence level p = the proportion of the target population

d = margin of error (5%, if the confidence level is 95%); 0.05, q = 1 - p; (1 - 0.5=0.5).

With a 10% non-response rate, the final sample size for the study was 422.

### Sampling technique and procedure

After conveniently selecting four public hospitals in the zone including Wallaga University referral hospital, Nekemte Comprehensive Specialized Hospital, Sire Primary Hospital, and Arjo Primary Hospitals as a study area and the sample sizes were allocated to the four respective hospitals based on their previous three-month delivery load based on the available data (report) from July 8, 2021 to July 7, 2022. After calculating the required sample size by using the single population proportion formula, sample size among the selected public hospitals was taken by proportionating for each hospital based on the total quarterly delivery load and required sample size. A systematic random sampling technique was used to interview the mothers in respective public hospitals until the allocated sample size was achieved. Generally, the interview with every five selected mothers from all the mothers attending public hospitals to utilize institutional delivery service during the data collection period was continued until the required sample size was reached. The sampling procedure for women who gave birth at public hospitals in the East Wallaga zone is depicted schematically (Fig 1).

**Fig 1:**
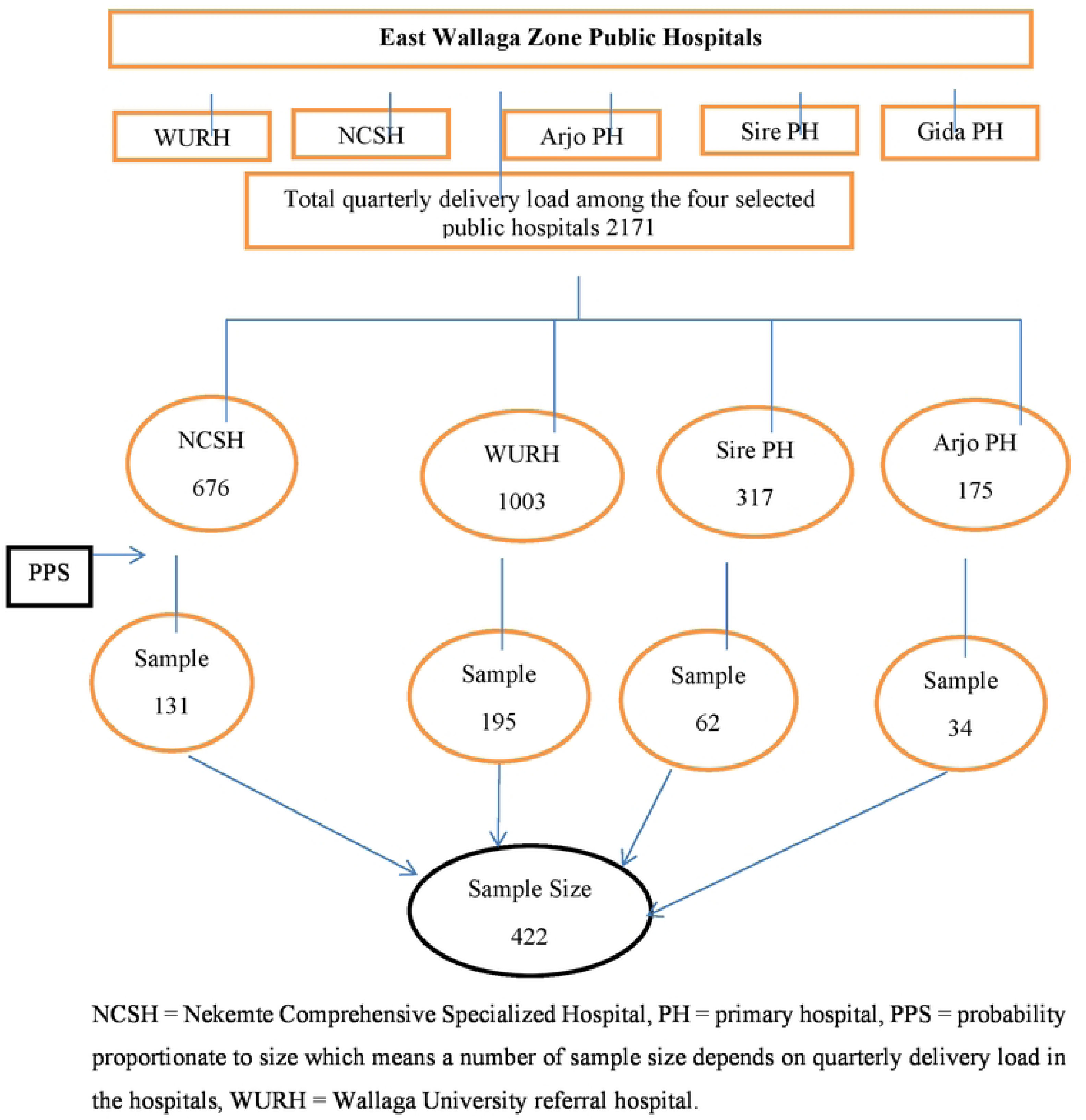
Schematic presentation of the sampling procedure for women who gave birth at public hospitals in East Wallaga zone, Oromia, Ethiopia.

### Study variables

**Dependent variable: –** Delays in utilization of institutional delivery.

**Independent variables: –** Socio-economic/demographic factors such as maternal age, educational status of the mother, educational status of the husband, decision-making power, place of residence, monthly income, mother’s and husband’s occupation. Obstetric-related factors included parity, antenatal care (ANC) follow-up, ANC frequency, and history of chronic illness during pregnancy, maternal knowledge of obstetric danger signs, and birth preparedness and complication readiness. Accessibility of the health facility such as accessibility of facility considering distance, cost of transportation, transportation availability, and accessibility to roads. Health system-related factors include staff availability, supplies, equipment, and waiting time to utilize delivery care. Individual level factors such as knowledge, attitude, and concern about privacy, perception of risks of labor, and delivery, and values on the benefits of institutional delivery. Cultural factors, myths, and rumors around childbirth including birth are natural and should take place at home, a strange environment can hurt the newborn, and the newborn may be exposed to air draft when delivered at health facilities.

### Data collection instruments and procedures

One of the research instruments, a questionnaire to collect quantitative data was adapted from similar studies, and JHPIEGO tools and indicators for monitoring maternal and neonatal health (16) and prepared in English and translated into Afan Oromo version for better understanding of data collectors and the study participants and then translated Afan Oromo version was translated back to English by consulting university English teachers for better grammar and coherence of the sentences and to ensure consistency, with one trained data collector assigned to each hospital. The quantitative data was collected using a standardized pre-tested questionnaire designed in such a way that it includes all the relevant variables to meet the study objectives. The questionnaire was designed to have four parts: socio-demographic characteristics of the study participants, accessibility to health services, women’s obstetric history, and antenatal care and delivery service utilization.

### Data quality control

Data quality was strictly maintained by carefully preparing data collection tools and creating a common consensus for data collectors through detailed training. Data quality may be affected at different points including questionnaire designing, data collection, and data entry unless measures are taken. As this is one of the points to control the quality of data, due emphasis was given to questionnaire designing. Objective-based, logically sequenced, free of scientific terms, and non-leading structured questionnaire was prepared. The data collectors and supervisors were provided with training on the objective of the study, the contents of the questionnaires, and how to maintain the confidentiality and privacy of the study participants. For clarity, the questionnaire was pre-tested outside the hospital in the selected health center one week before the actual data collection by using 5% of the sample size, and necessary corrections such as the sequencing of questions in a questionnaire were made depending on the ambiguity experienced in a pilot study. The collected data was checked by the principal investigator for completeness daily. The data were double-entered to verify the missing values.

### Data processing and analysis

The collected data were checked for completeness, edited, coded, and entered into Epi-data version 4.6, and exported to SPSS 24 for both descriptive and inferential statistical analysis with both binary and multivariate logistic regressions done to determine the association between dependent and independent variables. The data were summarized with descriptive statistics being computed for all variables according to type. Frequency, mean and standard deviation were obtained for continuous variables while the categorical variables were assessed by computing frequencies. Bivariate analysis with a 95% confidence interval was used to infer associations. In the initial step, bivariate analysis is employed to see the association between explanatory and outcome variables. Controlling the effect of confounding factors, each variable was entered into the logistic regression model as the independent variable with a dependent variable. Then, variables that were found statistically significant under bivariate analysis with p-value < 0.25 were entered into multiple logistic regression models to identify independent predictors of delays to utilize institutional delivery service and multicollinearity was checked by Hosmer and Lemeshow’s goodness of fit model. Finally, the adjusted odd ratio (AOR) along with a 95% confidence interval (CI) calculated for each candidate variable of interest to measure the strength of association and level of statistical significance was declared at p-value < 0.05.

### Operational definition

We defined delays according to the three-delay model (17). **Delays in utilization of institutional delivery service**: time taken more than one hour to decide to seek care or more than one hour to reach the health facility after making the decision or waiting for more than one hour in a health facility to receive delivery care. **First delay**: the time interval from the first onset of labor to the decision to seek emergency obstetric care. **Second delay**: delay to go to a health facility after the decision has been made to seek emergency obstetric care. There is a delay in physically reaching the nearby care facility within an hour after deciding to seek healthcare. **Third delay**: refers to receiving appropriate care once present at the health facility within the first 60 minutes of arrival. **Institutional delivery service utilization**: when a mother gives birth at a health institution and delivery is assisted by skilled birth attendants. Institutional delivery is interchangeably used with facility-based delivery and skilled birth attendance. **Women who gave birth at public hospitals**: women who are attended by health professionals at the hospital at the time of delivery during the data collection period. **Good knowledge of danger signs of labor**: knowing appropriately 3 or more danger signs or correctly replying to 3 well-known danger signs among the danger signs of labor. **Good knowledge of danger signs of pregnancy**: knowing appropriately 3 or more danger signs or correctly replying to 3 well-known danger signs among the danger signs of pregnancy. **Public hospitals**: Government hospitals that provide delivery service as one of any health institution.

### Ethical consideration

This study has been performed following the Belmont Report and Declaration of Helsinki. First, ethical clearance was obtained from the Research and Ethics Committee of Wallaga University; Ref no: WU/RD/622; Date: 24/01/2023. Then, letters of permission were sought from the East Wallaga Zonal Health Office. Following approval of ethical clearance and permission, formal letters were secured from the East Wallaga Zonal Health Office to each of the selected public hospitals. After being permitted to conduct the research at the selected hospitals, informed consent was obtained from each respondent before the actual data collection. Each respondent was informed about the objective of the study. The participants were told that their participation was purely voluntary and that their rights to not respond at all were respected and their confidentiality strictly maintained.

## Results

### Respondents’ socio-demographic characteristics

A face-to-face interview was held with systematically selected 422 mothers who gave birth at the selected public hospitals in the East Wallaga zone with a 100% response rate. Among the respondents, 283 (67.1%) of mothers were urban dwellers. The mean age of the respondents was 28.57 ± 4.8. Regarding their educational status, 203 (48.1%) of them attained secondary education and above (**Table 1**).

**Table 1:** Socio-demographic characteristics of women (n=422) who gave birth at selected public hospitals in East Wallaga zone, Oromia, Ethiopia, 2023.

| Variables | Frequency |  |
| --- | --- | --- |
|  | Number | Percent |
| **Place of Residence** |  |  |
| Urban | 283 | 67.1 |
| Rural | 139 | 32.9 |
| **Age in years** |  |  |
| 15-24 | 70 | 16.6 |
| 25-29 | 180 | 42.7 |
| ≥ 30 | 172 | 40.8 |
| **Maternal education** |  |  |
| Cannot read and write | 34 | 8.1 |
| Completed primary education | 185 | 43.8 |
| Completed secondary education and above | 203 | 48.1 |
| **Husband's Education** |  |  |
| Cannot read and write | 15 | 3.6 |
| Completed primary education | 134 | 31.7 |
| Completed secondary education and above | 273 | 64.7 |
| **Religion** |  |  |
| Orthodox | 104 | 24.6 |
| Protestant | 261 | 61.8 |
| Muslim | 49 | 11.6 |
| **Maternal occupation** |  |  |
| Unemployed | 338 | 80.1 |
| Employed | 84 | 19.9 |
| **Monthly family income on average** |  |  |
| > 1000 ETB | 393 | 86.6 |
| ≤ 1000 ETB | 29 | 13.4 |

### Women’s obstetric-related factors

The mean age of the mothers’ marriage was 21.97 ± 2.77 and the mean age of their first pregnancy was 23.66 ± 2.89. The majority (94.1%) and (93.6%) of them had ANC follow-up and information about the benefits of institutional delivery, respectively. More than one-fourth (25.8%) of them had maternal complications during their current pregnancy (**Table 2**).

**Table 2:** Obstetric characteristics of women (n=422) who gave birth at selected public hospitals in East Wallaga zone, Oromia, Ethiopia, 2023.

| Variables | Frequency |  |
| --- | --- | --- |
|  | Number | percent |
| **Age at first marriage** |  |  |
| 15-19 | 67 | 15.9 |
| 20-24 | 281 | 66.6 |
| 25-34 | 74 | 17.5 |
| **Age at first pregnancy** |  |  |
| 15-24 | 261 | 61.8 |
| 25-35 | 161 | 38.2 |
| **Gravida** |  |  |
| 1 | 117 | 27.7 |
| $\geq 2$ | 305 | 72.3 |
| **Parity** |  |  |
| 1 | 141 | 33.4 |
| ≥ 2 | 281 | 66.6 |
| **ANC follow-up** |  |  |
| Yes | 397 | 94.1 |
| No | 25 | 5.9 |
| **Number of ANC follow-up** |  |  |
| 1 | 22 | 5.2 |
| ≥ 2 | 375 | 88.9 |
| **Have information about the benefits of institutional delivery** |  |  |
| Yes | 395 | 93.6 |
| No | 27 | 6.4 |
| **If yes, the main source of information** |  |  |
| Healthcare providers | 348 | 82.5 |
| Mass media | 47 | 11.1 |
| **Knowledge about pregnancy health risks** |  |  |
| Yes | 302 | 71.6 |
| No | 120 | 28.4 |
| **Counseled about the danger signs of pregnancy** |  |  |
| Yes | 339 | 80.3 |
| No | 83 | 19.7 |
| **Having been told about birth plan/preparedness** |  |  |
| Yes | 391 | 92.7 |
| No | 31 | 7.3 |
| **Knowledge about danger signs of labor** |  |  |
| Yes | 325 | 77.0 |
| No | 97 | 23.0 |
| **Who decided to seek care?** |  |  |
| My self | 382 | 90.5 |
| Husband | 40 | 9.5 |
| **Previous history of obstetric complications** |  |  |
| Yes | 109 | 25.8 |
| No | 313 | 74.2 |
| **Maternal complication during a previous pregnancy** |  |  |
| Yes | 97 | 23.0 |
| No | 325 | 77.0 |
| **Maternal complications during current pregnancy** |  |  |
| Yes | 109 | 25.8 |
| No | 313 | 74.2 |

### Health services accessibility-related factors

Among the women interviewed, 166 (39.3%) of the mothers lived more than 10 kilometers from the health facility, and 173 (41.0%) of the mothers encountered transportation problems. The reasons for transportation problems were living in areas that are inaccessible to any transport services, with only dry weather roads, and the cost of transport was expensive (**Table 3**).

**Table 3:** Health services accessibility-related factors among women who gave birth at selected public hospitals in East Wallaga zone, Oromia, Ethiopia, 2023.

| variables | Frequency |  |
| --- | --- | --- |
|  | Number | Percent |
| **Distance in kilometers from the nearest health center** |  |  |
| <10 | 256 | 60.7 |
| >10 | 166 | 39.3 |
| **Distance in kilometers from the nearest health post** |  |  |
| <10 | 291 | 69.0 |
| >10 | 131 | 31.0 |
| **Having money for transport when labor started** |  |  |
| Yes | 368 | 87.2 |
| No | 54 | 12.8 |
| **Transportation problems on the way to health facilities** |  |  |
| Yes | 173 | 41.0 |
| No | 249 | 59.0 |
| **If yes, reasons for transportation problem** |  |  |
| Inaccessible to any transport service | 118 | 28.0 |
| Only dry weather road | 22 | 5.2 |
| Expensive | 33 | 7.8 |
| **Means of transport to reach the nearest health center** |  |  |
| Motorized transport | 307 | 72.7 |
| Animal | 22 | 5.2 |
| Walking | 70 | 16.6 |
| People support carrying the mother | 23 | 5.5 |

### Health system-related factors

Among the mothers interviewed, 201 (47.6%) of them were referral cases. Most (87.9%) of the referral cases were referred once whereas 10.1% of them were referred twice among the referral cases and the majority (93.5%) the referred from the health center. Among all, 410 (97.2%) of them prefer to attend a health institution for delivery (**Table 4**).

**Table 4:** Health system-related factors associated with institutional delivery utilization among women who gave birth at selected public hospitals in East Wallaga zone, Oromia, Ethiopia, 2023.

| Variables | Frequencies |  |
| --- | --- | --- |
|  | Number | percent |
| **Referral case** |  |  |
| Yes | 199 | 47.2 |
| No | 223 | 52.8 |
| **If yes, the number of referral** |  |  |
| Once | 175 | 87.9 |
| Twice | 20 | 10.1 |
| More than twice | 4 | 2.0 |
| **If yes, reasons for referral** |  |  |
| Lack of trained staff | 1 | 0.5 |
| Lack of drugs/equipment/supplies | 93 | 46.7 |
| Absent staff | 2 | 1.0 |
| Obstetric complications | 103 | 51.8 |
| **If yes, referring unit** |  |  |
| Private clinic | 9 | 4.5 |
| Health center | 186 | 93.5 |
| Hospital | 4 | 2.0 |
| **Preference to attend health facility for delivery** |  |  |
| Yes | 410 | 97.2 |
| No | 12 | 2.8 |
| **Delay to seek care to be a factor affecting pregnancy outcome** |  |  |
| Yes | 411 | 97.4 |
| No | 11 | 2.6 |
| **A traditional issue that causes women to delay utilizing delivery services** |  |  |
| Yes | 402 | 95.3 |
| No | 20 | 4.7 |

### The magnitude of maternal delays

This study revealed that the magnitude of the first maternal delay was 47.2% [95% CI: (42.7, 51.7)] and the mean delay time was 0.47 ± 0.5 hours. The magnitude of the second maternal delay was 71.3% [95% CI: (64.5, 78.1)]. The mean delay time for the second delay was 0.71 hours with a standard deviation of 0.45 hours. The magnitude of the third maternal delay was 10.2% [95% CI: (9.23, 11.17)]. The mean third maternal delay time was 1.9 with a standard deviation of 0.3. The magnitude of the total maternal delays was 82.5% (Fig 2).

**Fig 2:**
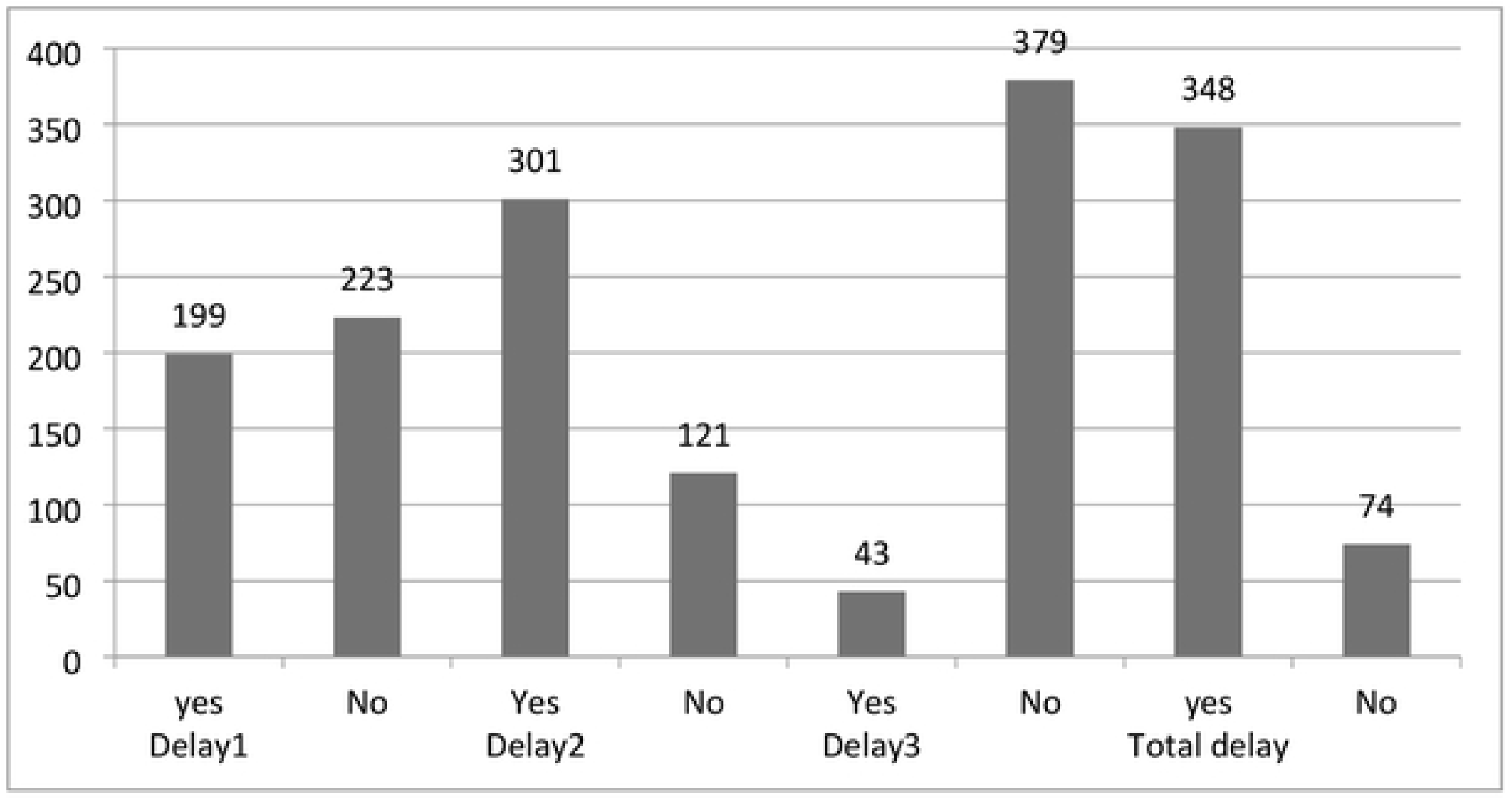
Maternal, delays encountered in utilization of institutional, delivery among women who gave birth at selected public hospitals (n=422) in East Wallaga zone Oromia, Ethiopia, 2023.

### Factors associated with first maternal delay

Information on institutional delivery, decision maker to seek care, knowledge about danger signs of labor, counseling on danger signs of pregnancy, counseling on a birth plan, complications in a previous pregnancy, complications in current pregnancy, distance from health center, referral case, transportation problem, and availability of money for transport were found to be significantly associated with the first delay in binary logistic regression analysis. After controlling for the effects of confounding variables, two variables: complications in current pregnancy, and referral were found to be factors significantly associated with first maternal delay in multiple logistic regression analysis.

Accordingly, the odds of the first delay were higher (1.73 times) [AOR = 1.73, 95% CI: (1.08, 2.76)] among mothers who had complications in the current pregnancy than their counterparts. The odds of the first delay were higher (1.60 times) [AOR = 1.60, 95% CI: (1.04, 2.47)] among mothers who experienced referral from one facility to another facility than mothers who utilized institutional delivery at the first contact of visit (**Table 5**).

**Table 5:**
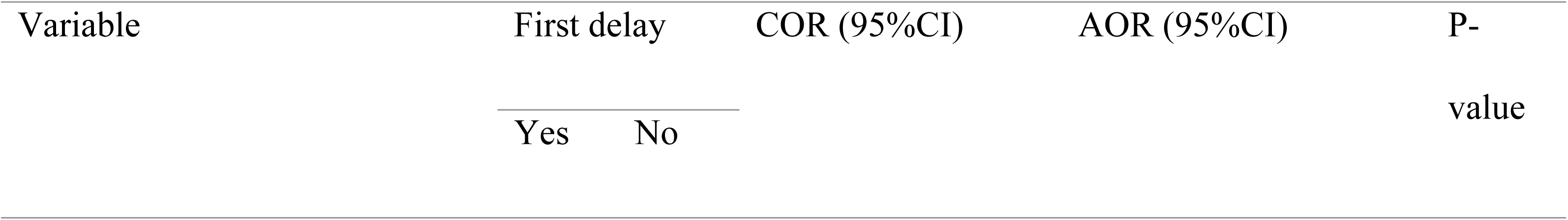
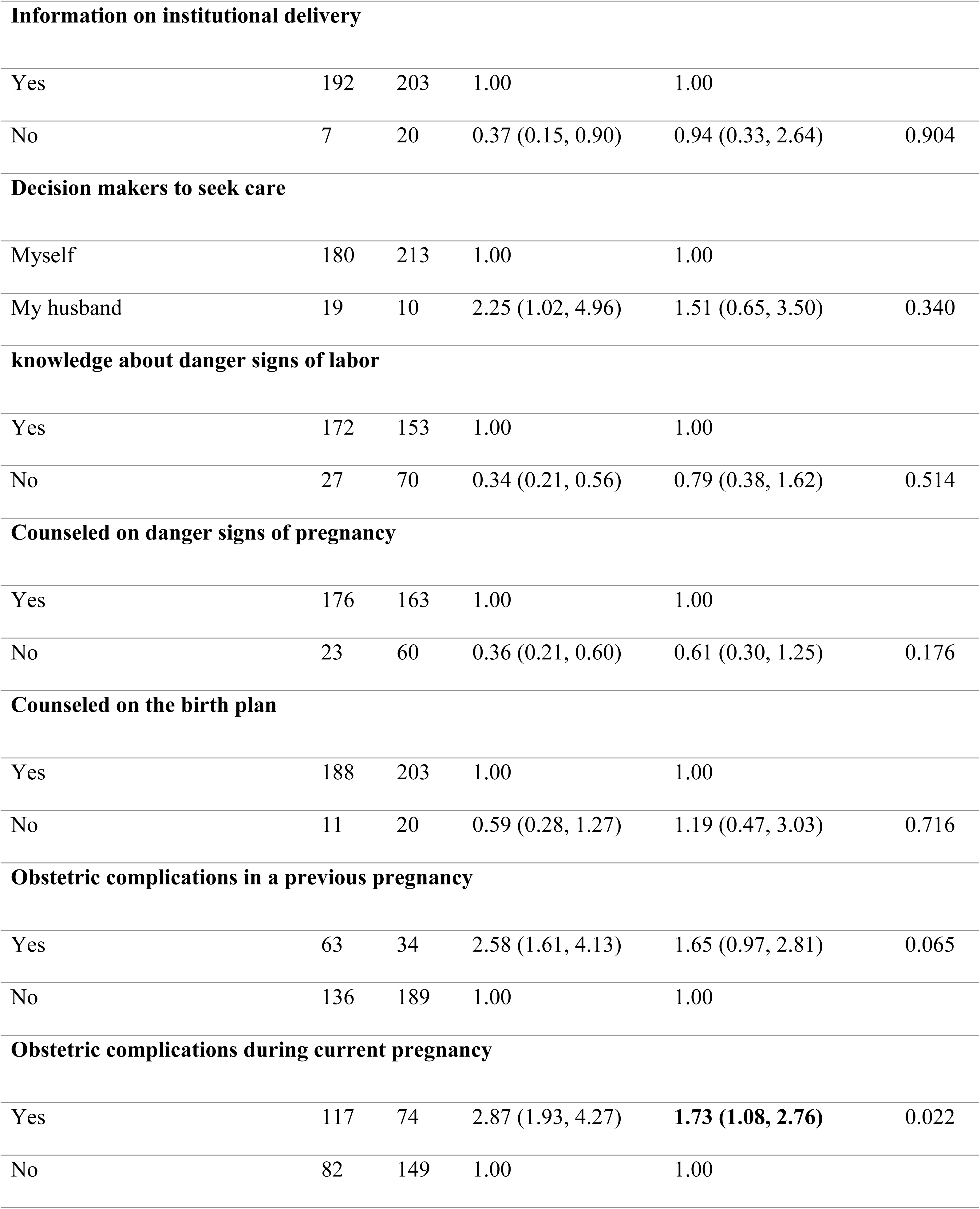
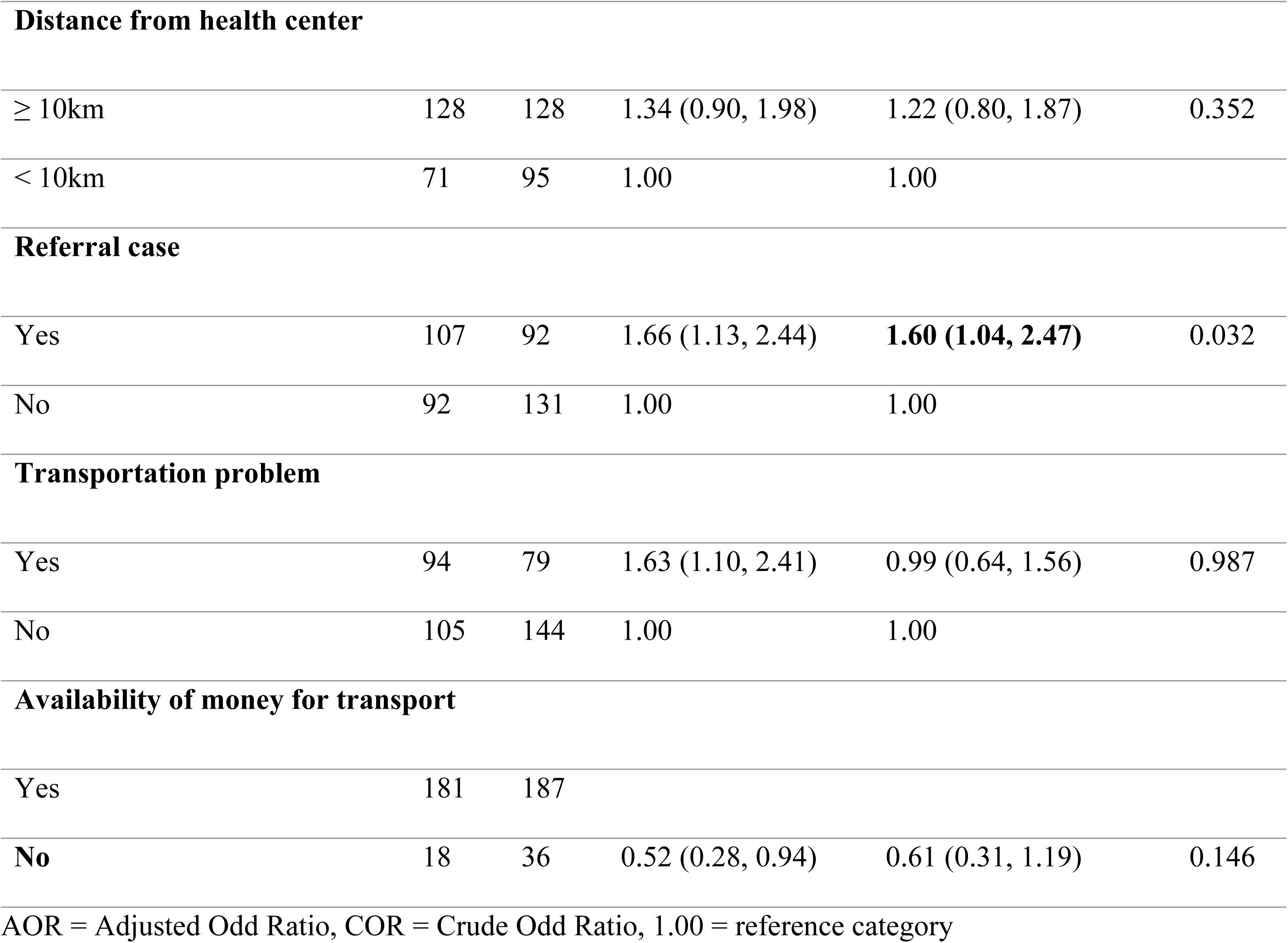
Multivariable regression results for factors associated with the first maternal delay among women who gave birth at public hospitals in East Wallaga Zone, Oromia, Ethiopia, 2023.

### Factors associated with second maternal delay

Information on institutional delivery, knowledge about danger signs of labor, counseling on danger signs of pregnancy, complications in the current pregnancy, referral case, and availability of money for transport were found to be candidate variables for multivariable logistic regression with the second delay. After controlling for the effects of confounding variables, knowledge about danger signs of labor was independently associated with second maternal delay in multiple logistic regression analysis. Accordingly, mothers who did not know the danger signs of labor had 2.93 times higher odds [AOR = 2.93, 95% CI: (1.47, 5.86)] of the second maternal delay than their counterparts (**Table 6**).

**Table 6:**
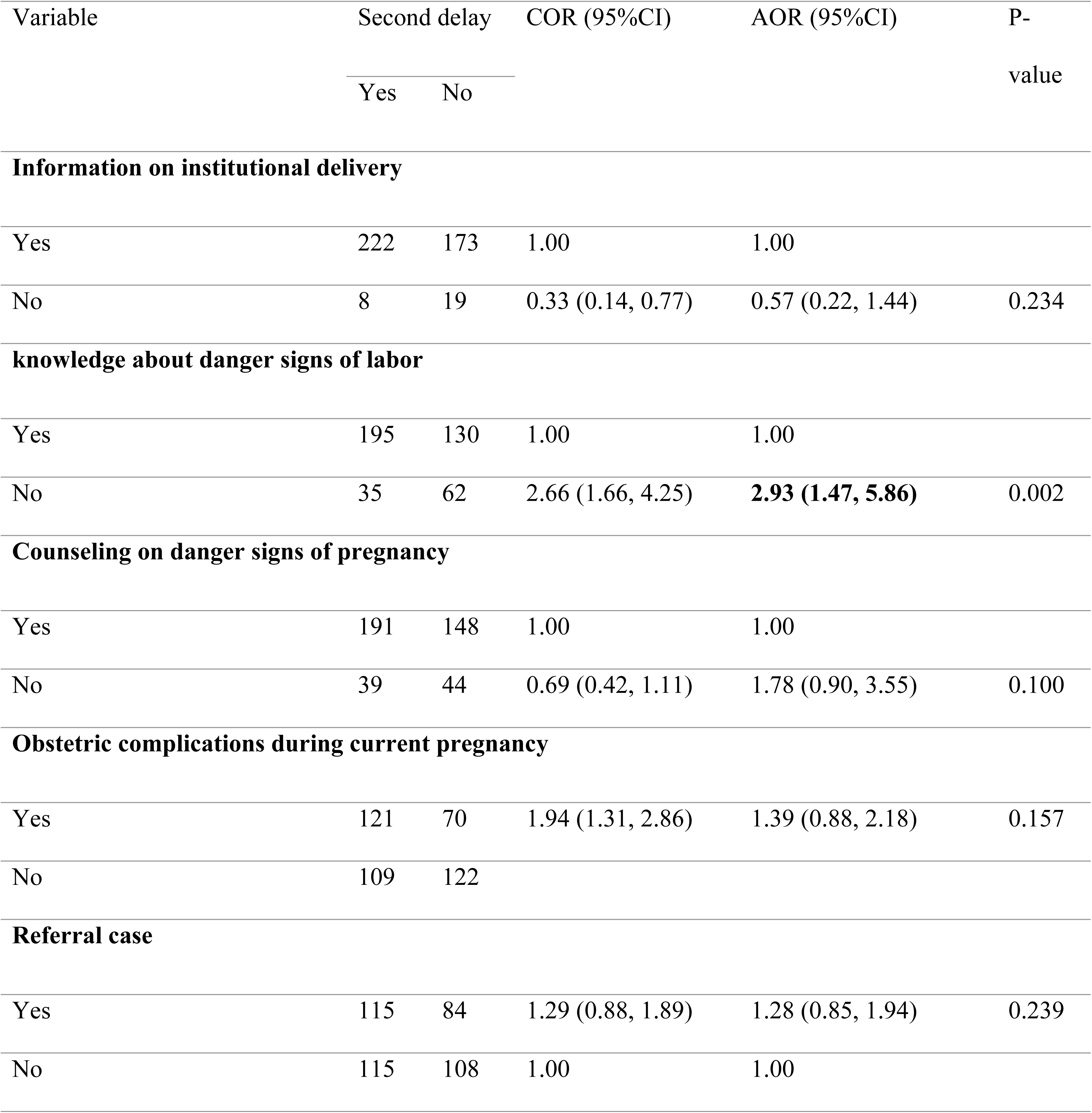
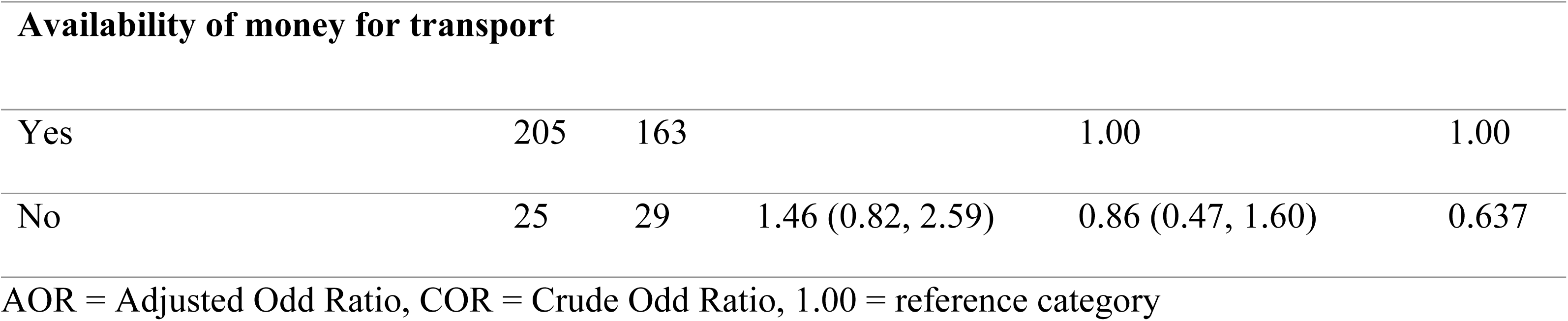
Multivariable regression results for factors associated with the second maternal delay among women who gave birth at public hospitals in East Wallaga Zone, Oromia, Ethiopia, 2023.

| Variable | Second delay |  | COR (95%CI) | AOR (95%CI) | P-value |
| --- | --- | --- | --- | --- | --- |
|  | Yes | No |  |  |  |
| Information on institutional delivery |  |  |  |  |  |
| Yes | 222 | 173 | 1.00 | 1.00 |  |
| No | 8 | 19 | 0.33 (0.14, 0.77) | 0.57 (0.22, 1.44) | 0.234 |
| knowledge about danger signs of labor |  |  |  |  |  |
| Yes | 195 | 130 | 1.00 | 1.00 |  |
| No | 35 | 62 | 2.66 (1.66, 4.25) | 2.93 (1.47, 5.86) | 0.002 |
| Counseling on danger signs of pregnancy |  |  |  |  |  |
| Yes | 191 | 148 | 1.00 | 1.00 |  |
| No | 39 | 44 | 0.69 (0.42, 1.11) | 1.78 (0.90, 3.55) | 0.100 |
| Obstetric complications during current pregnancy |  |  |  |  |  |
| Yes | 121 | 70 | 1.94 (1.31, 2.86) | 1.39 (0.88, 2.18) | 0.157 |
| No | 109 | 122 |  |  |  |
| Referral case |  |  |  |  |  |
| Yes | 115 | 84 | 1.29 (0.88, 1.89) | 1.28 (0.85, 1.94) | 0.239 |
| No | 115 | 108 | 1.00 | 1.00 |  |
| **Availability of money for transport** |  |  |  |  |  |
| Yes | 205 | 163 |  | 1.00 | 1.00 |
| No | 25 | 29 | 1.46 (0.82, 2.59) | 0.86 (0.47, 1.60) | 0.637 |

AOR = Adjusted Odd Ratio, COR = Crude Odd Ratio, 1.00 = reference category

### Factors associated with the third maternal delay

Awareness of health risks in pregnancy, knowledge about danger signs of labor, counseling on danger signs of pregnancy, complications in current pregnancy, decision maker to seek care, referral, transport problem, and availability of money for transport were found to be significantly associated with the third delay in binary logistic regression analysis. After controlling for the effects of confounding variables, two variables: complications in current pregnancy and availability of money for transport were found to be factors significantly associated with third maternal delay in multiple logistic regression analysis.

Accordingly, the odds of the third delay were higher (more than two times) [AOR = 2.26, 95% CI: (1.01, 5.05)] among mothers who had obstetric complications in their current pregnancy than their counterparts. Similarly, the odds of the third delay were higher (nearly four times) [AOR = 3.98, 95% CI: (1.66, 9.57)] among mothers who had no money for transport than their counterparts (**Table 7**).

**Table 7:** Multivariable regression results for factors associated with the third maternal delay among women who gave birth at public hospitals in East Wallaga Zone, Oromia, Ethiopia, 2023.

| Variable | Third delay |  | COR (95%CI) | AOR (95%CI) | P-value |
| --- | --- | --- | --- | --- | --- |
|  | Yes | No |  |  |  |
| Awareness of health risks in pregnancy |  |  |  |  |  |
| Yes | 36 | 266 | 1.00 | 1.00 |  |
| No | 7 | 113 | 2.19 (0.94, 5.06) | 0.57 (0.21, 1.56) | 0.273 |
| knowledge about danger signs of labor |  |  |  |  |  |
| Yes | 38 | 287 | 1.00 | 1.00 |  |
| No | 5 | 92 | 0.41 (0.16, 1.07) | 0.62 (0.19, 1.99) | 0.423 |
| Obstetric complications during current pregnancy |  |  |  |  |  |
| Yes | 29 | 162 | 2.78 (1.42, 5.42) | 2.26 (1.01, 5.05) | 0.048 |
| No | 14 | 217 | 1.00 | 1.00 |  |
| Decision makers to seek care |  |  |  |  |  |
| Myself | 37 | 356 | 1.00 | 1.00 |  |
| My husband | 6 | 23 | 2.51 (0.96, 6.56) | 1.41 (0.49, 4.10) | 0.523 |
| Referral case |  |  |  |  |  |
| Yes | 26 | 173 | 1.82 (0.96, 3.47) | 1.65 (0.84, 3.23) | 0.148 |
| No | 17 | 206 | 1.00 | 1.00 |  |

| Transportation problem |  |  |  |  |  |
| --- | --- | --- | --- | --- | --- |
| Yes | 19 | 154 | 2.69 (1.26, 5.72) | 0.93 (0.46, 1.87) | 0.827 |
| No | 24 | 225 | 1.00 | 1.00 |  |
| Availability of money for transport |  |  |  |  |  |
| Yes | 32 | 336 | 1.00 | 1.00 |  |
| No | 11 | 43 | 0.52 (0.28, 0.94) | **3.98 (1.66, 9.57)** | 0.002 |

AOR = Adjusted Odd Ratio, COR = Crude Odd Ratio, 1.00 = reference category

## Discussion

In the current study, we assessed the magnitude and associated factors of first, second, and third delays in utilizing institutional delivery among the study participants. Accordingly, the magnitude of the first maternal delay was 47.2%. This finding is comparable with studies in Jimma Medical Center (46.7%), Southern Ethiopia (44.2%), and Addis Ababa (51.7%) (18–20). However, it exceeds the findings from studies in Bahir Dar (37.8%), and Hadiya zone (40.1%) (21, 22). The possible reason for this discrepancy might be due to the differences in sample sizes, the time gap of the study, variations in the distribution of health facilities, and social, demographic, and geographic variations in the study area.

In this study, the magnitude of the second maternal delay was 71.3%. This finding is in line with the study done in Melbourne (74%) (23) and Pakistan (74%) (24). However, the findings of the current study are lower than a study in Afghanistan (80%) (25). The possible reason for the difference may be due to geographic, demographic, and methodological variations between the studies. However, the current finding is higher than the findings of studies in Southern Ethiopia (43.2%) (20), Bangladesh (38%) (26), Bahir Dar (31.7%) (21), Hadiya zone (29.7%) (22), Afghanistan (65%) (25), and Addis Ababa (44.8%) (19). The difference might be due to the inaccessibility of health facilities, place of study settings, unavailability of roads and inadequate ambulance service, and the economic status of women. The possible explanation for the difference may also be due to the socio-demographic, geographic and cultural variation, sample size, and distribution of the health infrastructures among the study areas. This implies that many mothers spend their time in an attempt to reach the health facilities (23).

The magnitude of the third maternal delay was 10.2% which is comparable with the study conducted in Burkina Faso (8.2%) (27) and Mozambique (14.2%) (28). This indicates there is good handling of cases in the hospitals once the mothers reached there attributable to well-equipped and staffed hospitals, especially those specialized and referral hospitals. However, this result is lower than the studies in Sidama Regional State (29.3%) (29), Gamo Zone (31.7%) (30), Arsi Zone (25.5%) (31), Bahir Dar (30.7%) (21), Amhara Regional State (26.9%) (32), Southwest Ethiopia (34.7%) (20), Gurage Zone (34.8%) (33), and Addis Ababa (58.6% (19). This difference might be due to variations in the study setting, sample size, population size, women’s socioeconomic and demographic characteristics, fee-free services in the study area, advanced medical logistics supply, and professional staff. For example, the two referral hospitals in Addis Ababa provide services with fees, and the population size was incomparably high in contrast to the study area which may affect service provision.

In this study, the first maternal delay was associated with obstetric complications during the current pregnancy. Accordingly, mothers who had obstetric complications during their current pregnancy had a 1.73 times higher first delay in the decision to seek to utilize institutional delivery than their counterparts. This finding is supported by a study done in Southern Ethiopia (33). The possible explanation might be due to pregnant women who had no antenatal care visit could not get more information about the risks and complications they may encounter during pregnancy (34).

Mothers referred from other healthcare facilities were 1.70 times more likely to experience the first maternal delay in utilizing institutional delivery than non-referred mothers (a woman who was not referred from another healthcare facility and gave birth at the selected hospital). This finding is supported by the studies conducted in Mozambique, the South Gondar zone of Amhara Regional State, Hadiya zone, and Oromia regional state (28, 35–37). The possible reason might be due to inadequate financial and logistical support, a severe shortage of skilled human resources for health, and an ineffective referral mechanism (38).

This study found mothers who had poor knowledge of danger signs of labor had 2.93 times higher odds of second maternal delay as compared to those who had good knowledge. This finding is supported by the studies done in the Gamo zone of Southern Ethiopia (39) and the Bale zones of the Oromia region (40). The possible explanation might be due to women’s poor awareness of obstetric danger signs during labor and delivery (41).

The current study revealed that complications of current pregnancy were a statistically significant factor with a third maternal delay. Accordingly, those women who encountered obstetric complications in their current pregnancy had 2.26 times higher odds of experiencing a third maternal delay in utilizing institutional delivery service compared to their counterparts and this finding is supported by a study done in Gurage zone, Southern Ethiopia (33) and Southeastern Ethiopia (40). The possible justification for not timely utilizing institutional delivery might be due to a lack of skilled professionals, equipment, drugs, and supplies at the health facilities, and the presence of complications that might need additional diagnostic procedures, which can contribute to longer wait times for receiving care (42).

In this study, women who had no money for transport had almost four times higher odds of experiencing a third maternal delay in utilizing institutional delivery service compared to their counterparts. This finding is comparable with a study done in Bahir Dar City of Amhara Regional State (21). This might be due to the socio-economic inequalities, inability to pay for the expensive private car rent at inconvenient times, and poor readiness of the family members to accompany the mothers to the health facilities (43). This could be also due to lack of money being one of the major factors for home delivery and shaping the choice of delivery place. Availability of money for transport was noted as an important factor in whether health facility delivery is sought and the main obstacles to accessing the hospital for emergency obstetric care were lack of money and transportation difficulties (44).

## Limitations of the study

The current study has some limitations which will be addressed by future researchers. This study was conducted at public hospitals, and the laboring mothers are not representative of the East Wallaga Zone’s general population. Recall bias may have influenced the results since the mothers were interviewed shortly after giving delivery when they were emotionally and physically spent. Therefore, future researchers should conduct a community-based investigation to address the current study’s limitations.

## Conclusion

As indicated by the study, there was a high magnitude of first and second-maternal delays in utilizing institutional delivery in the study area. However, the magnitude of third maternal delays in the utilization of institutional delivery services was relatively lower than in the previous studies. The current history of obstetric complications in pregnancy, referral cases, knowledge of the danger signs of labor, and availability of money for transport were statistically associated with maternal delays in utilizing institutional delivery. To further reduce delays, this study emphasizes the significance of addressing three delays in seeking institutional delivery services. Not only the availability of community and government support are necessary to help the mothers get rid of the delays but also maternal attitudes towards utilization of institutional delivery are very important to attend institutional delivery early reducing the delays.

## Supporting information

S1 File. English version of questionnaire, information sheet and consent form. (DOCX) S2 File. Data set. (Sav)

## Data Availability

All relevant data are within the manuscript and its supporting information files.

## Acknowledgment

We would like to express our sincere gratitude and deep appreciation to the Research and Ethics Committee of Wallaga University for the approval of ethical clearance. We would also like to extend our gratitude to the East Wallaga Zonal Health Department for their letter of permission to conduct the study. Finally, our special thanks go to the data collectors, supervisors, and participants.

## Authors’ contributions

**Conceptualization:** Getahun Tulu Amante, Gemechu Dereje Feyissa.

**Data curation:** Getahun Tulu Amante, Gemechu Dereje Feyissa.

**Formal analysis:** Getahun Tulu Amante, Gemechu Dereje Feyissa, Markos Desalegn, Emiru Merdassa.

**Investigation:** Getahun Tulu Amante.

**Methodology:** Getahun Tulu Amante, Gemechu Dereje Feyissa, Markos Desalegn, Emiru Merdassa.

**Project administration:** Getahun Tulu Amante, Gemechu Dereje Feyissa.

**Resources:** Getahun Tulu Amante, Gemechu Dereje Feyissa, Mokonnen Dereje.

**Software:** Getahun Tulu Amante, Gemechu Dereje Feyissa, Mokonnen Dereje.

**Supervision:** Getahun Tulu Amante, Markos Desalegn, Emiru Merdassa.

**Validation:** Getahun Tulu Amante, Gemechu Dereje Feyissa, Markos Desalegn, Emiru Merdassa.

**Visualization:** Getahun Tulu Amante, Mokonnen Dereje, Markos Desalegn, Emiru Merdassa.

**Writing – original draft:** Getahun Tulu Amante, Gemechu Dereje Feyissa.

**Writing – review & editing:** Gemechu Dereje Feyissa.

## Consent for publication

Not applicable.

## Abbreviations

AOR: Adjusted odds ratio

CI: Confidence interval

COR: Crude odds ratio

CSA: Central Statistical Agency

EDHS: Ethiopian Demographic and Health Survey

HSTP: Health sector transformation plan

MMR: Maternal mortality ratio

SDGs: Sustainable development goals

SPSS: Statistical package for social science

WHO: World health organization

